# Efficacy of therapeutic plasma exchange for the treatment of autoimmune hemolytic anemia: Protocol for a meta-analysis of randomized controlled trials

**DOI:** 10.1101/2020.02.03.20020214

**Authors:** Jiawen Deng

## Abstract

Autoimmune hemolytic anemia (AIHA) is a rare blood disorder that results in hemolysis of red blood cells (RBCs) due to the presence of autoantibodies in the serum. Previous research has shown that the use of therapeutic plasma exchange therapy (TPE) may be effective at treating AIHA by removing autoimmune antibodies from the intravascular space. However, very little knowledge synthesis is available on the use of TPE in AIHA patients due to the rarity of the disease. We propose a meta-analysis that investigates whether the use of TPE, with or without concurrent treatment regimens, can decrease adverse events, increase remission rate and improve lab results including hemoglobin, RBC, reticulocyte counts, hematocrit and total bilirubin.

## INTRODUCTION

Autoimmune hemolytic anemia (AIHA) is a rare blood disorder characterized by the hemolysis of self red blood cells (RBCs) as a result of the production of autoantibodies[1]. Previous research had shown that the use of therapeutic plasma exchange (TPE) may be effective in increasing remission in AIHA patients[1]. However, there are no knowledge synthesis in this area of study, since AIHA is a rare disease. We propose to conduct a systematic review and meta-analysis to investigate whether the use of TPE can decrease adverse events, increase the rate of remission and improve lab figures.

## METHODS

We will conduct this meta-analysis in accordance to the Preferred Reporting Items for Systematic Reviews and Meta-Analyses (PRISMA) framework[2]. This study is currently being reviewed for registration on The International Prospective Register of Systematic Reviews (PROSPERO). Any significant amendments to this protocol will be reported and published with the results of the review.

This study is conducted concurrently with the project “Therapeutic use of blood products for the treatment of autoimmune hemolytic anemia: A network meta-analysis”, and thus they share the same search strategy and various aspects of study design. The protocol for the “Therapeutic use of blood products for the treatment of autoimmune hemolytic anemia: A network meta-analysis” can be found here: https://doi.org/10.1101/2020.01.15.20017657.

### Eligibility Criteria

#### Types of Participants

We will include adult patients (18 years or older) who have been diagnosed with autoimmune hemolytic anemia, defined as per individual study criteria.

#### Types of Interventions

We will first include all studies that include the use of TPE. We will then categorize the included studies using the following classifications, as reported by ter Veer et al[3]:

1. Treatment A vs. Treatment B
2. Treatment B+A vs. Treatment B+C
3. Treatment A vs. Treatment A+B

Studies satisfying category 3), where Treatment B consists of TPE, will be included. The concurrent treatment arm, Treatment A, should remain the same between treatment arms. Studies where TPE is compared to no treatment will be included as well. If a study contains more than 2 treatment arms, only the treatment arms satisfying category 3) will be included.

#### Types of Studies

We will include randomized and quasi-randomized parallel-groups RCTs.

### Primary Outcomes

#### Remission Incidence (n)

We will evaluate incidence of remission based on data collected at the latest follow-up. Definitions of remission will be defined as per individual study criteria. We expect the definitions of remission to be a combination of improvements in clinical symptoms and lab results.

### Secondary Outcomes

#### Lab Results

We will evaluate hemoglobin count (g/L), RBC count (10^12^/L), reticulocyte count (%), hematocrit (%), and total bilirubin (μmol/L) based on the latest lab results.

#### Adverse Events (n)

We will evaluate the incidence of adverse events based on data collected at the latest follow-up. Definitions of adverse events will be defined as per individual study criteria.

### Search Methods for Identification of Studies

#### Electronic Database Search

We will conduct a database search of MEDLINE, EMBASE, Web of Science, CINAHL, and CENTRAL from inception to January 2020. We will use relevant MeSH headings to ensure appropriate inclusion of titles and abstracts (see ***Table 1*** for search strategy).

**Table 1.** MEDLINE Search Strategy

| Database: OVID Medline Epub Ahead of Print, In-Process & Other Non-Indexed Citations, Ovid MEDLINE(R) Daily and Ovid MEDLINE(R) 1946 to Present |  |
| --- | --- |
| Line |  |
| 1 | exp Anemia, Hemolytic, Autoimmune/ |
| 2 | ((Cold or Hot or Warm) adj2 Agglutinin Disease?).ti,ab,kw,kf. |
| 3 | ((Cold or Hot or Warm) adj2 Antibody Disease?).ti,ab,kw,kf. |
| 4 | ((Cold or Hot or Warm) adj2 Antibody H?emolytic Anemia?).ti,a... |
| 5 | Acquired Autoimmune H?emolytic Anemia.ti,ab,kw,kf. |
| 6 | Idiopathic Autoimmune H?emolytic Anemia.ti,ab,kw,kf. |
| 7 | Secondary Autoimmune H?emolytic Anemia.ti,ab,kw,kf. |
| 8 | Autoimmune H?emolytic Anemia/ |
| 9 | AIHA.ti,ab,kw,kf. |
| 10 | WAIHA.ti,ab,kw,kf. |
| 11 | or/1-10 |
| 12 | exp randomized controlled trial/ |
| 13 | exp Randomized Controlled Trials as Topic/ |
| 14 | random*.mp. |
| 15 | Random Allocation/ |
| 16 | ((singl* or doubl* or tripl* or trebl*) adj3 (blind* or mask*)).mp. |
| 17 | double-blind method/ or single-blind method/ |
| 18 | or/12-17 |
| 19 | 11 and 18 |

Major Chinese databases, including Wanfang Data, Wanfang Med Online, CNKI, and CQVIP will also be searched using a custom Chinese search strategy.

The study strategy utilized in this study will be shared with the study “Therapeutic use of blood products for the treatment of autoimmune hemolytic anemia: A network meta-analysis”.

#### Other Data Sources

We will also conduct hand search the reference list of previous meta-analyses and NMAs for included articles.

### Data Collection and Analysis

#### Study Selection

We will perform title and abstract screening independently and in duplicate using Rayyan QCRI (https://rayyan.qcri.org). Studies will only be selected for full-text screening if both reviewers deem the study relevant. Full-text screening will also be conducted in duplicate. We will resolve any conflicts via discussion and consensus or by recruiting a third author for arbitration.

#### Data Collection

We will carry out data collection independently and in duplicate using data extraction sheets developed a priori. We will resolve discrepancies by recruiting a third author to review the data.

#### Risk of Bias

We will assess risk of bias (RoB) independently and in duplicate using The Cochrane Collaboration’s tool for assessing risk of bias in randomized trials[4]. Two reviewers will assess biases within each article in seven domains: random sequence generation, allocation concealment, blinding of participants and personnel, blinding of outcome assessment, incomplete outcome data, selective reporting, and other biases (see ***Table 2*** for definitions of RoB domains).

**Table 2.**
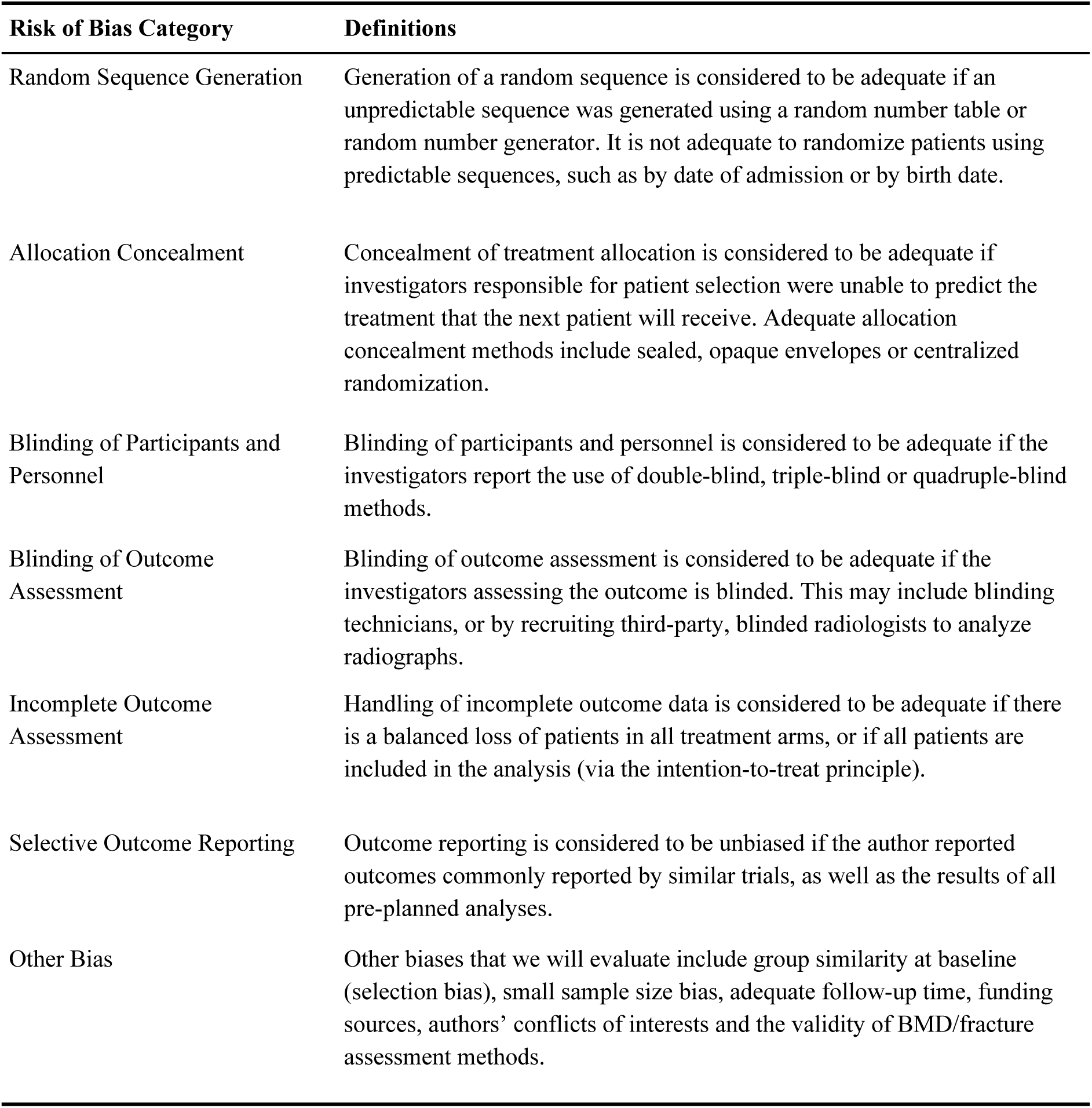
Definitions of Risk of Bias Domains

If a majority of domains are considered to be low risk, the study will be assigned a low RoB. Similarly, if a majority of domains are considered to be high risk, the study will be assigned a high RoB. If more than half of the domains have unclear risk or if there are an equivalent number of low and high, low and unclear or high and unclear domains, the study will be assigned an unclear RoB.

### Data Items

#### Bibliometric Data

Authors, year of publication, trial registration, digital object identifier (DOI), publication journal, funding sources and conflict of interest.

#### Methodology

\# of participating centers, study setting, blinding methods, phase of study, enrollment duration, randomization and allocation methods, criteria for remission.

#### Baseline Data

\# randomized, # analyzed, mean age, sex, baseline lab results, follow up duration.

#### Outcomes

\# of patients in remission at the latest follow up, lab results at the latest follow up, # of patients who had experienced at least one adverse event at the latest follow up.

### Statistical Analysis

#### Meta-Analysis

We will conduct all statistical analyses using R 3.6.2[5]. We will perform meta-analyses using the *meta* library. Because we expect significant heterogeneity among studies due to differences in methodology, we will use a random effects model[6].

For remission incidence and adverse event incidence we will report the results of the analyses as risk ratio (RR), pooled using the Mantel-Haenszel method[7], with 95% confidence intervals (CIs) with Hartung-Knapp adjustment for random effects model[8]. For Hb, RBC, reticulocyte counts, hematocrit and total bilirubin, we will report the results as mean differences (MDs), pooled using the inverse variance method[9], with corresponding 95% CIs with Hartung-Knapp adjustment for random effects model[8]. We used the Sidik-Jonkman estimator[8,9] for τ^2^ calculations, and we used the Q-profile method[10] for estimating the confidence interval of τ^2^ and τ.

#### Missing Data

We will attempt to contact the authors of the original studies to obtain missing or unpublished data. If we cannot obtain missing standard deviations (SDs), the study will be excluded from the analysis even if the mean was provided. SDs will not be imputed.

#### Heterogeneity Assessment

We will assess statistical heterogeneity using I^2^ statistics, τ^2^ and Cochran’s Q[11]. We will identify the source of heterogeneity by

a. identify outlier studies with treatment effect that is not included in the 95% confidence interval of the pooled effect size using the *dmetar* library;
b. perform influence analyses, or the “leave-one-out” analysis, where the meta-analysis is repeated with one study omitted, using the *dmetar* library;
c. Perform GOSH analyses[12] using the *metafor* library. The GOSH plot will be examined for evaluable clusters, and *gosh.diagnostics* function will be used to identify the outlying studies if there are evaluable clusters.

We will perform sensitivity analyses excluding outlier studies to observe the outliers’ effects on the original pooled effect size and heterogeneity measures.

#### Publication Bias

We will use funnel plots[13] to detect the presence of small study effects. We will use Egger’s test[14] to check for asymmetry within the funnel plot to identify possible publication bias. If Egger’s test reveals significant publication bias, we will use the trim-and-fill method[15] to estimate the actual effect size with imputations of the missing small studies. This will be done using the *trimfill* method in the *meta* library.

We will also perform p-curve[16] analyses to detect the presence of “p-hacking”[17] using the *dmetar* library. We will report whether we observed evidence of “p-hacking”, such as a lack of right skew in the p-curve plot or low estimated statistical power.

#### Meta-Regression

We will perform meta-regression on:

1. % of patients with primary/secondary AIHA
2. % of patients with warm/cold AIHA
3. % female patients
4. follow up periods

## Data Availability

No data is available for this manuscript.

## ACKNOWLEDGEMENTS

None

## AUTHOR STATEMENT

JD made significant contributions to conception and design of the work, drafted the work, and substantially reviewed it.

## FUNDING

This research received no specific grant from any funding agency in the public, commercial or not-for-profit sectors.

## CONFLICTS OF INTEREST

No potential conflicts of interest were reported by the authors.

## Notes

### Competing Interest Statement

The authors have declared no competing interest.

### Funding Statement

No funding was received for this study.

## REFERENCES

1 Silberstein LE, Berkman EM. Plasma exchange in autoimmune hemolytic anemia (AIHA). J Clin Apher 1983;1:238–42. doi:10.1002/jca.2920010407

2 Moher D, Liberati A, Tetzlaff J, et al. Preferred reporting items for systematic reviews and meta-analyses: the PRISMA statement. Ann Intern Med 2009;151:264–9, W64. doi:10.7326/0003-4819-151-4-200908180-00135

3 Ter Veer E, van Oijen MGH, van Laarhoven HWM. The Use of (Network) Meta-Analysis in Clinical Oncology. Front Oncol 2019;9:822. doi:10.3389/fonc.2019.00822

4 Higgins JPT, Altman DG, Gotzsche PC, et al. The Cochrane Collaboration’s tool for assessing risk of bias in randomised trials. BMJ. 2011;343:d5928-d5928. doi:10.1136/bmj.d5928

5 Team RC. R: A language and environment for statistical computing. 2015.

6 Serghiou S, Goodman SN. Random-Effects Meta-analysis: Summarizing Evidence With Caveats. JAMA 2019;321:301–2. doi:10.1001/jama.2018.19684

7 Mathes T, Kuss O. A comparison of methods for meta-analysis of a small number of studies with binary outcomes. Res Synth Methods 2018;9:366–81. doi:10.1002/jrsm.1296

8 IntHout J, Ioannidis JPA, Borm GF. The Hartung-Knapp-Sidik-Jonkman method for random effects meta-analysis is straightforward and considerably outperforms the standard DerSimonian-Laird method. BMC Med Res Methodol 2014;14:25. doi:10.1186/1471-2288-14-25

9 Marín-Martínez F, Sánchez-Meca J. Weighting by Inverse Variance or by Sample Size in Random-Effects Meta-Analysis. Educ Psychol Meas 2010; 70:56–73. doi: 10.1177/0013164409344534

10 Jackson D, Turner R, Rhodes K, et al. Methods for calculating confidence and credible intervals for the residual between-study variance in random effects meta-regression models. BMC Med Res Methodol 2014; 14:103. doi: 10.1186/1471-2288-14-103

11 Higgins JPT, Thompson SG, Deeks JJ, et al. Measuring inconsistency in meta-analyses. BMJ 2003; 327:557–60. doi: 10.1136/bmj.327.7414.557

12 Olkin I, Dahabreh IJ, Trikalinos TA. GOSH - a graphical display of study heterogeneity. Res Synth Methods 2012; 3:214–23. doi: 10.1002/jrsm.1053

13 Sterne JAC, Sutton AJ, Ioannidis JPA, et al. Recommendations for examining and interpreting funnel plot asymmetry in meta-analyses of randomised controlled trials. BMJ 2011; 343:d4002. doi: 10.1136/bmj.d4002

14 Lin L, Chu H. Quantifying publication bias in meta-analysis. Biometrics 2018; 74:785–94. doi: 10.1111/biom.12817

15 Duval S, Tweedie R. Trim and fill: A simple funnel-plot-based method of testing and adjusting for publication bias in meta-analysis. Biometrics 2000; 56:455–63. doi: 10.1111/j.0006-341x.2000.00455.x

16 Simonsohn U, Nelson LD, Simmons JP. P-curve won’t do your laundry, but it will distinguish replicable from non-replicable findings in observational research: Comment on Bruns & Ioannidis (2016). PLoS One. 2019; 14:e0213454. doi: 10.1371/journal.pone.0213454

17 Head ML, Holman L, Lanfear R, et al. The extent and consequences of p-hacking in science. PLoS Biol 2015; 13:e1002106. doi: 10.1371/journal.pbio.1002106

